# Generation of false positive SARS-CoV-2 antigen results with testing conditions outside manufacturer recommendations: A scientific approach to pandemic misinformation

**DOI:** 10.1101/2021.07.04.21259490

**Authors:** Glenn Patriquin, Ross J. Davidson, Todd F. Hatchette, Breanne M. Head, Edgard Mejia, Michael G. Becker, Adrienne Meyers, Paul Sandstrom, Jacob Hatchette, Ava Block, Nicole Smith, John Ross, Jason J. LeBlanc

## Abstract

**Objectives:** Antigen-based rapid diagnostics tests (Ag-RDTs) are useful tools for SARS-CoV-2 detection. However, misleading demonstrations of the Abbott Panbio COVID-19 Ag-RDT on social media claimed that SARS-CoV-2 antigen could be detected in municipal water and food products. To offer a scientific rebuttal to pandemic misinformation and disinformation, this study explored the impact of using the Panbio SARS-CoV-2 assay with conditions falling outside of manufacturer recommendations.

**Methods:** Using Panbio, various water and food products, laboratory buffers, and SARS-CoV-2-negative clinical specimens were tested, with and without manufacturer buffer. Additional experiments were conducted to assess the role of each Panbio buffer component (tricine, NaCl, pH, and tween-20), as well as the impact of temperatures (4°C, 20°C, and 45°C) and humidity (90%) on assay performance.

**Results:** Direct sample testing (without the kit buffer), resulted in false positive signals resembling those obtained with SARS-CoV-2-positive controls tested under proper conditions. The likely explanation of these artifacts is non-specific interactions between the SARS-CoV-2-specific conjugated and capture antibodies, as proteinase K treatment abrogated this phenomenon, and thermal shift assays showed pH-induced conformational changes under conditions promoting artifact formation. Omitting, altering, and reverse engineering the kit buffer all supported the importance of maintaining buffering capacity, ionic strength, and pH for accurate kit function. Interestingly, the Panbio assay could tolerate some extremes of temperature and humidity outside of manufacturer claims.

**Conclusions:** Our data support strict adherence to manufacturer instructions to avoid false positive SARS-CoV-2 Ag-RDT reactions, otherwise resulting in anxiety, overuse of public health resources, and dissemination of misinformation.

## Introduction

High demand for diagnostic testing during the COVID-19 pandemic led to the development of various technologies for SARS-CoV-2 detection.[1] Nucleic acid amplification tests (NAATs), like real-time RT-PCR, are considered the reference methods [1-3], but antigen-based rapid diagnostic tests (Ag-RDTs) have been widely used due to their ease-of use, rapid results, and ability to be performed outside of a laboratory setting.[1] Many Ag-RDTs have been licensed as point-of-care (POC) devices for SARS-CoV-2 detection [4,5], but their performance can vary between methods, testing frequency, and settings in which they are used.[6-12] Ag-RDTs are well recognized to be less sensitive and specific than commercial NAATs, and false positive results from Ag-RDTs are known to occur, particularly in settings of low disease prevalence.[13,14]

The intended use of the COVID-19 Ag Rapid Test Device is qualitative detection of SARS-CoV-2 antigen (i.e. nucleocapsid protein) from nasal swabs (or nasopharyngeal swabs, depending on the formulation of the kit). The manufacturer kit insert states that instructions must be strictly followed by a trained healthcare professional to achieve accurate results, and the kit includes a buffer used for antigen extraction from the swabs used for specimen collection as well as viral inactivation. However, misleading demonstrations of a SARS-CoV-2 Ag-RDT (i.e. Panbio) on social media platforms have claimed that SARS-CoV-2 antigen can readily be detected in municipal water and commercial food and beverages, if tested directly onto the Panbio test device.[15-18] Moreover, on social media, the misuse of an Ag-RDT was propagated by pupils in attempts to miss time in school.[15-18] However, in both these examples, the results are erroneous as direct testing of samples onto the Ag-RDTs device is not recommended by manufacturer. With misinformation and disinformation often perpetuated on social media, and aberrant results obtained from improper use of the kit, unsubstantiated claims can undermine confidence in SARS-CoV-2 diagnostic testing and erode trust in public health efforts. As such, it is important to use science-based approaches to demonstrate that while non-specific reactivity can occur when testing is performed under inappropriate conditions, SARS-CoV-2 is not truly present in food or potable water samples. For healthcare professionals, aberrant test results arising from procedures that deviate from the kit instructions would not be surprising. When manufacturer instructions are followed, the expected false positivity rate would be very low (i.e. between 0.4 and 1.2%) [6-12], and the positive Ag-RDTs are often repeated using an alternative method such as a NAAT.[1-3] On the other hand, the cause of false positive Ag-RDT reactions are rarely investigated.

This study deliberately evaluated conditions that fell outside of those recommended by the manufacturer, which had the potential to generate aberrant Ag-RDT reactions, including unregulated buffering capacity or ionic strength, and extremes of temperature, humidity, and pH. As expected from social media claims, direct testing of a wide variety of food products and water samples generated false positive results with the Panbio Ag-RDT; however, this prompted further investigations into the underlying mechanism of artifact generation. Panbio kit extraction buffer was omitted, diluted, or reverse engineered, to help demonstrate the importance of the buffer and each of its components. Overall, by identifying conditions that could favor artifact generation, this study not only helps provide evidence supporting the importance of following manufacturer instructions, but helps in the understanding of possible causes of false positive reactions using Ag-RDTs, which can be informative to healthcare professionals, test manufacturers, and other users of the products.

## Materials and methods

### Sample types

Ag-RDT samples included food products (Table 1), water, laboratory buffers, specimen transport media, and four different clinical specimens types previously tested negative by real-time RT-PCR in routine diagnostic testing: 1) 30 nasopharyngeal (NP) swabs in universal transport media (UTM); 2) 30 oropharyngeal and bilateral nares (OP/N) swabs in phosphate buffered saline (PBS) [19,20]; 3) 30 bronchoalveolar lavages (BAL); and 4) 30 saline gargles (Table 1).[21]

**Table 1.**
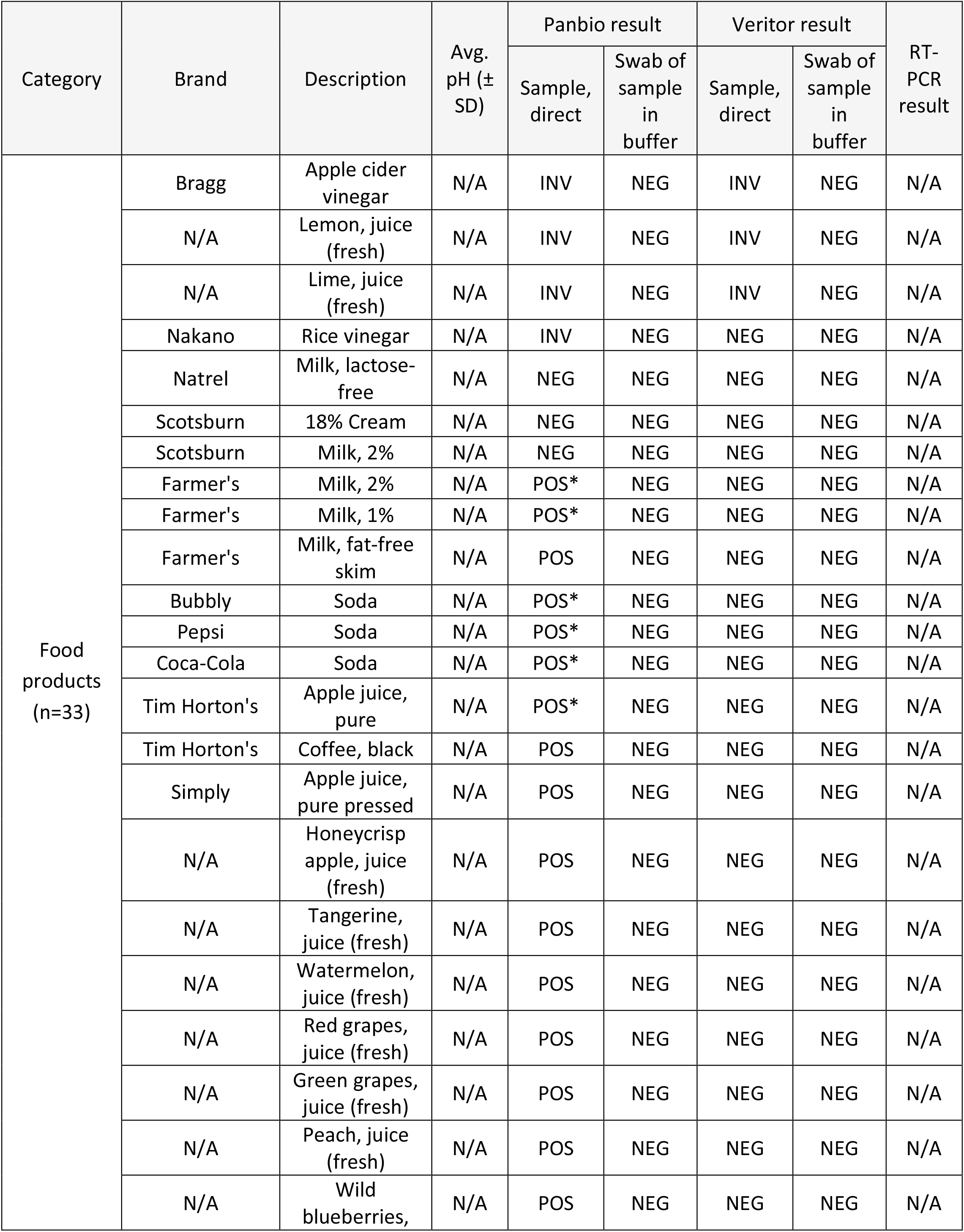
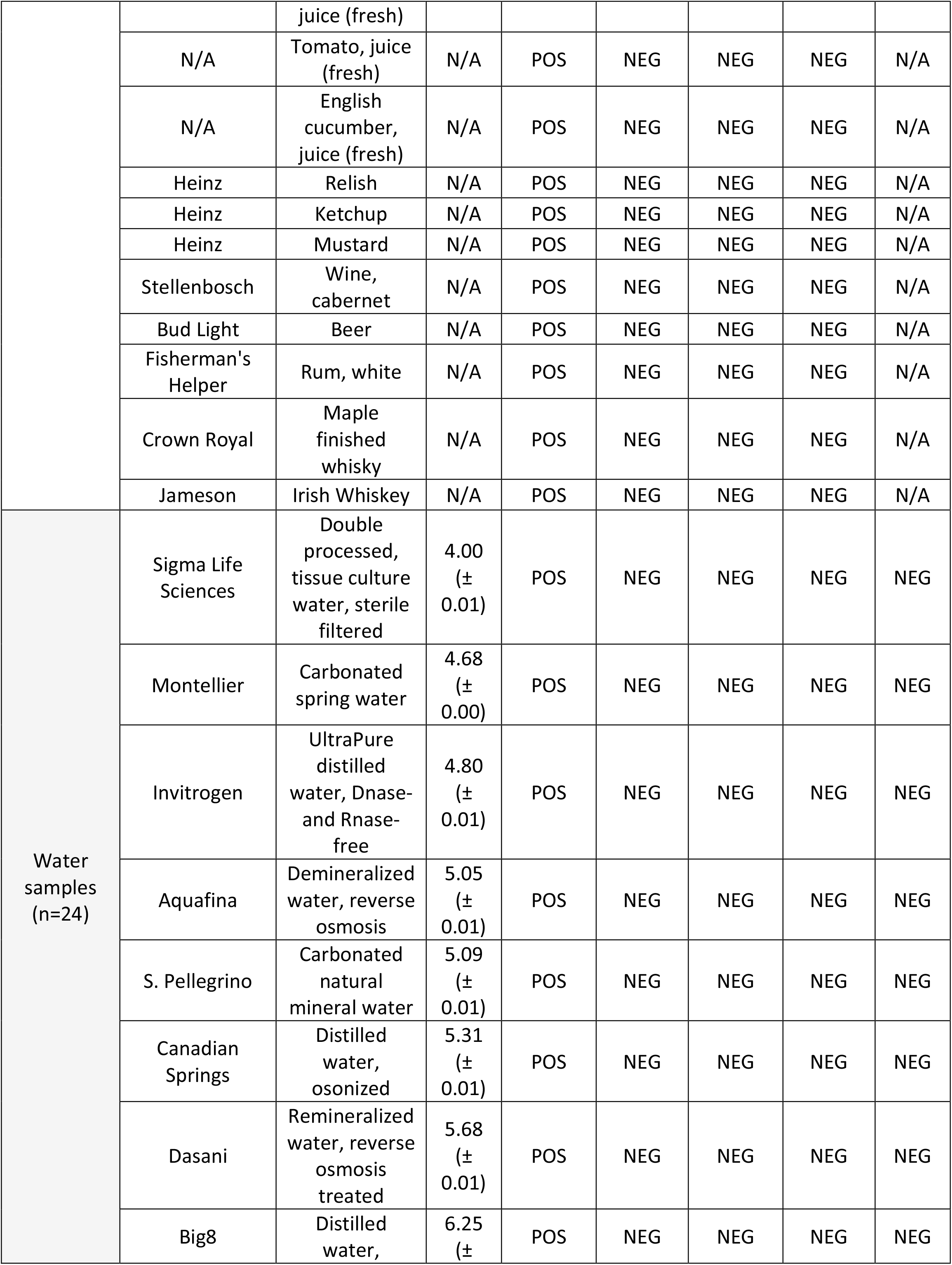
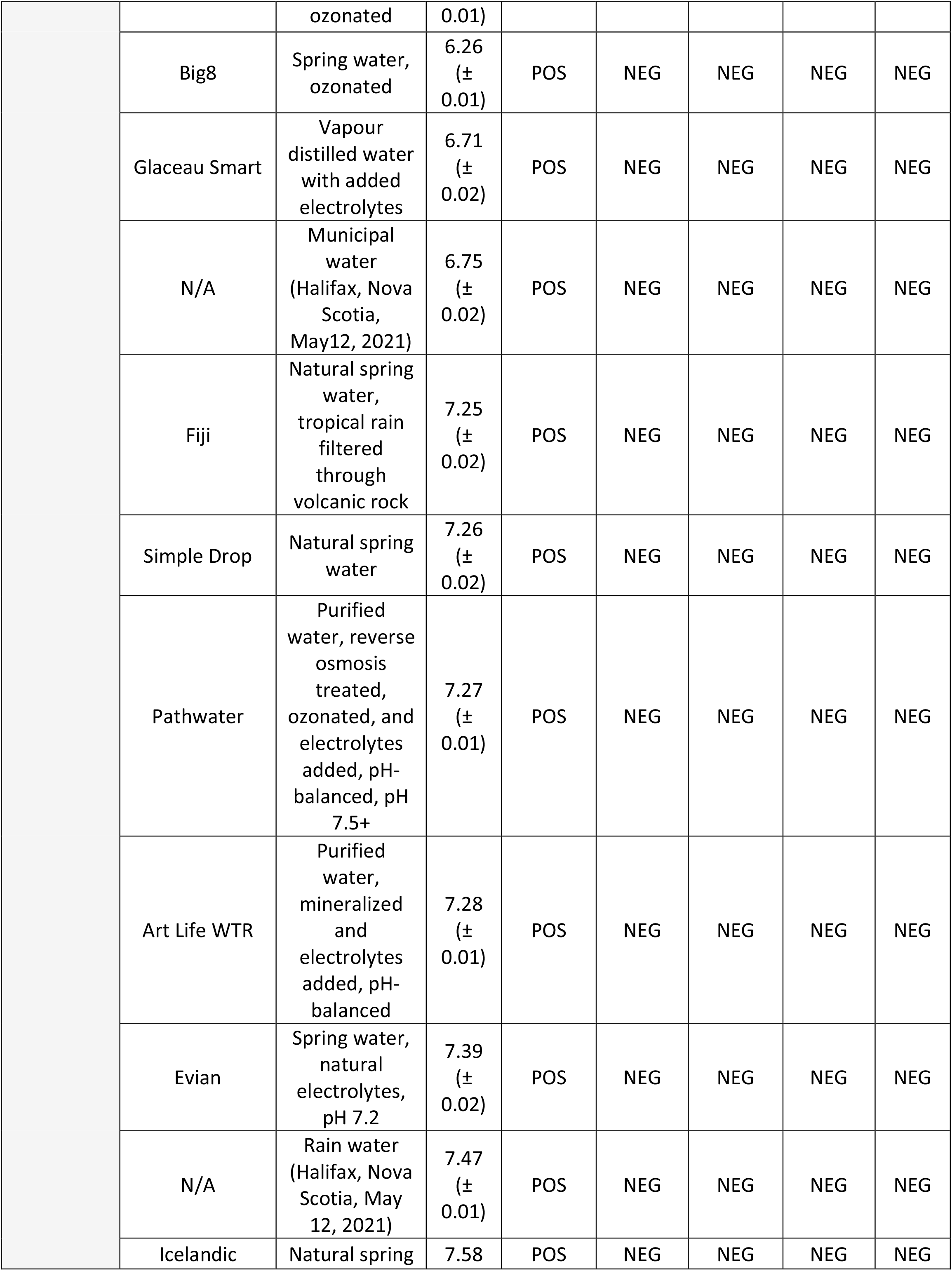
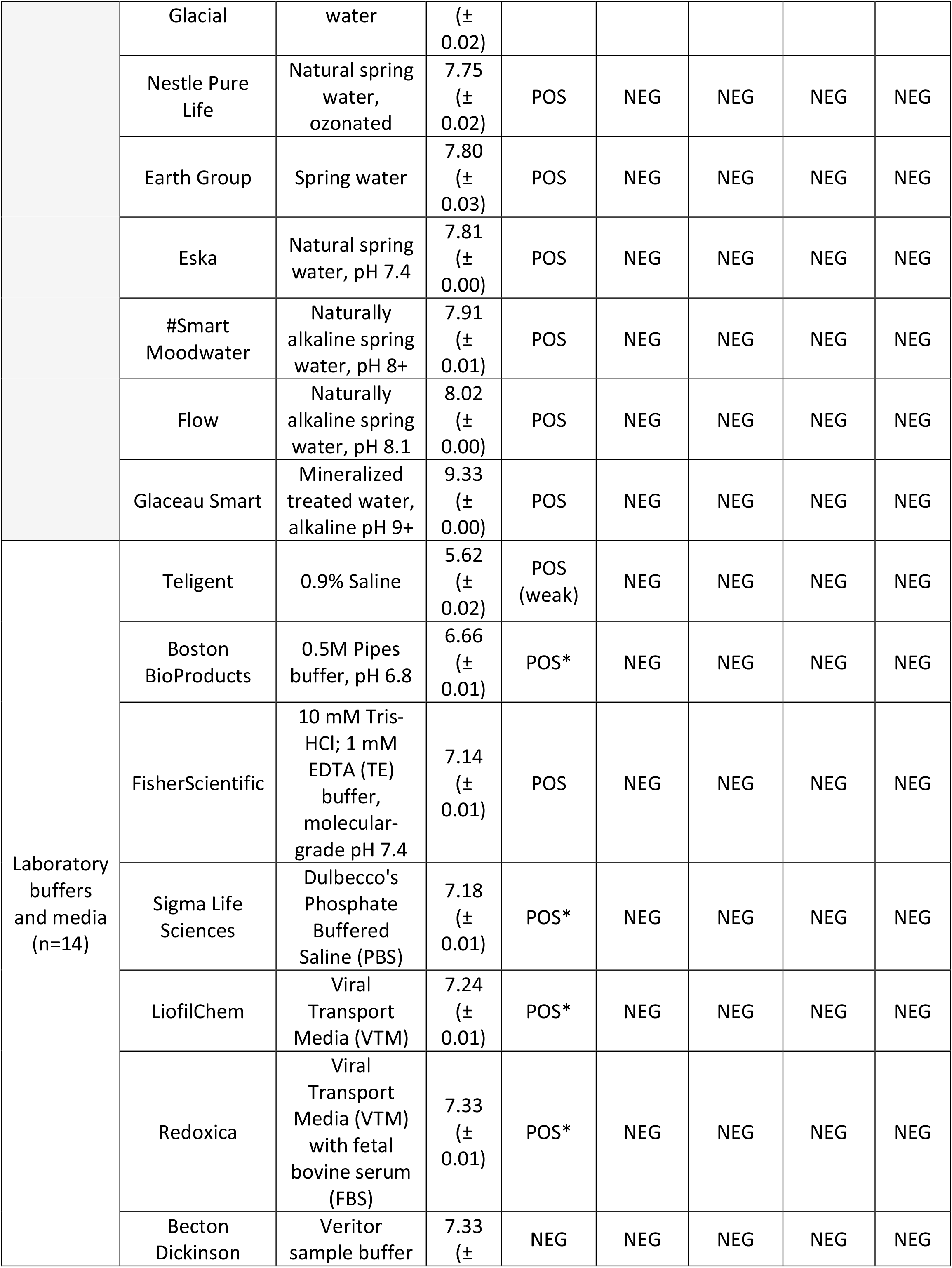
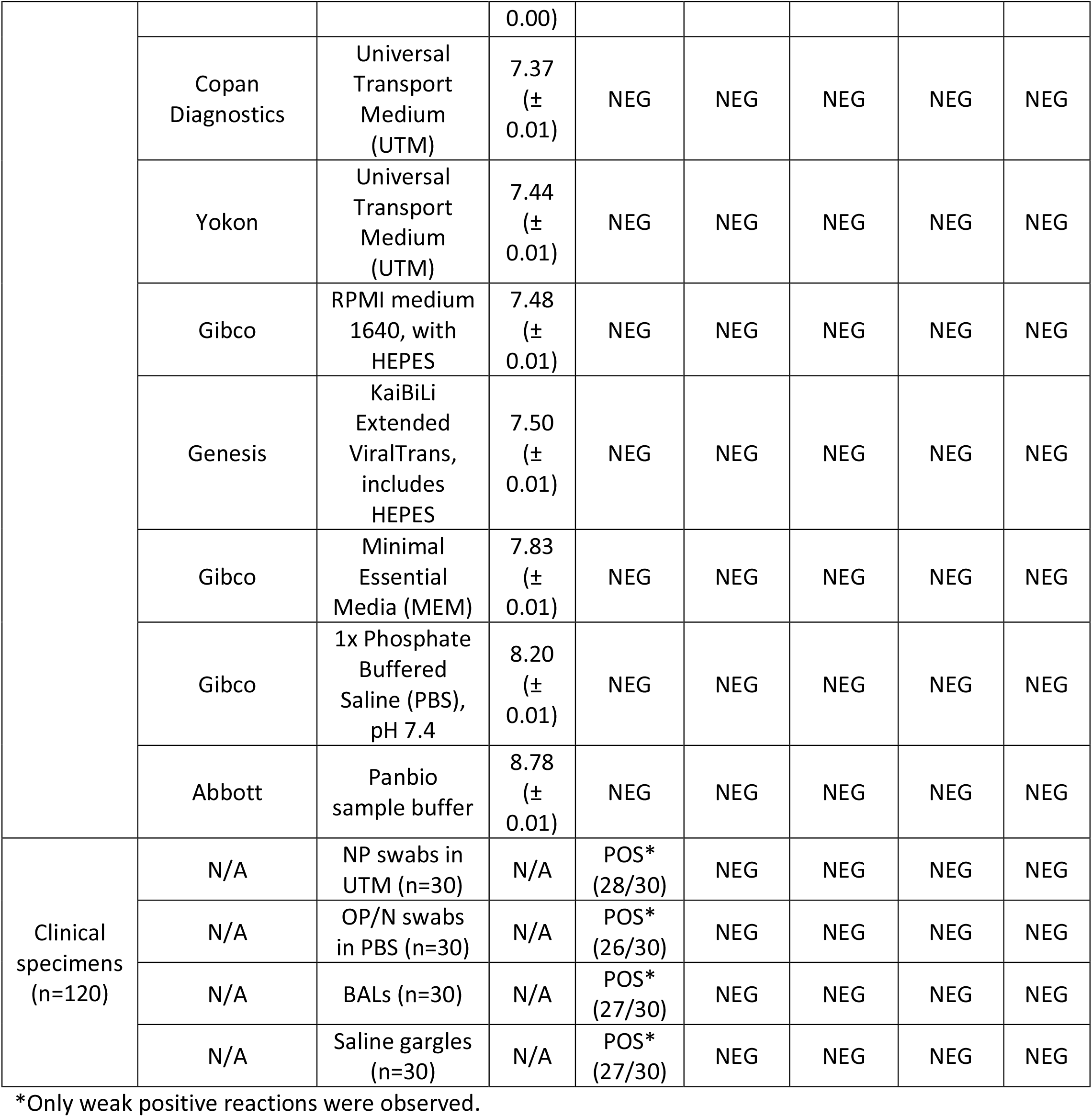
Samples tested by SARS-CoV-2 Ag-RDTs with and without manufacturer buffer.

### Antigen and molecular testing

SARS-CoV-2 nucleocapsid antigen detection was performed using the Abbott Panbio COVID-19 Rapid Antigen Test (Figure 1) and the BD Veritor System for Rapid Detection of SARS-CoV-2. Each kit’s nasal swabs were dipped into the test samples, placed in the appropriate kit buffers, and 3 or 5 drops were used to inoculate the sample wells of the Veritor and Panbio cassettes, respectively, as per the manufacturer recommendations. Each sample was also tested without manufacturer buffer (i.e. direct sample testing), mirroring the test procedure recommended for clinical specimens. Results were visualized by the unaided eye after 15 minutes, and Veritor readouts also included automated detection using a BD Veritor Plus instrument. Panbio test results were based on the kit insert, in which the presence of the control band alone was considered a negative results, the presence of both the control and target bands was considered positive, and the presence of the target band alone or absence of bands was considered invalid (Figure 1). Real-time RT-PCR testing was performed for all specimens except food products using the Roche Diagnostics cobas SARS-CoV-2 Test on the cobas 6800 instrument.

**Figure 1.**
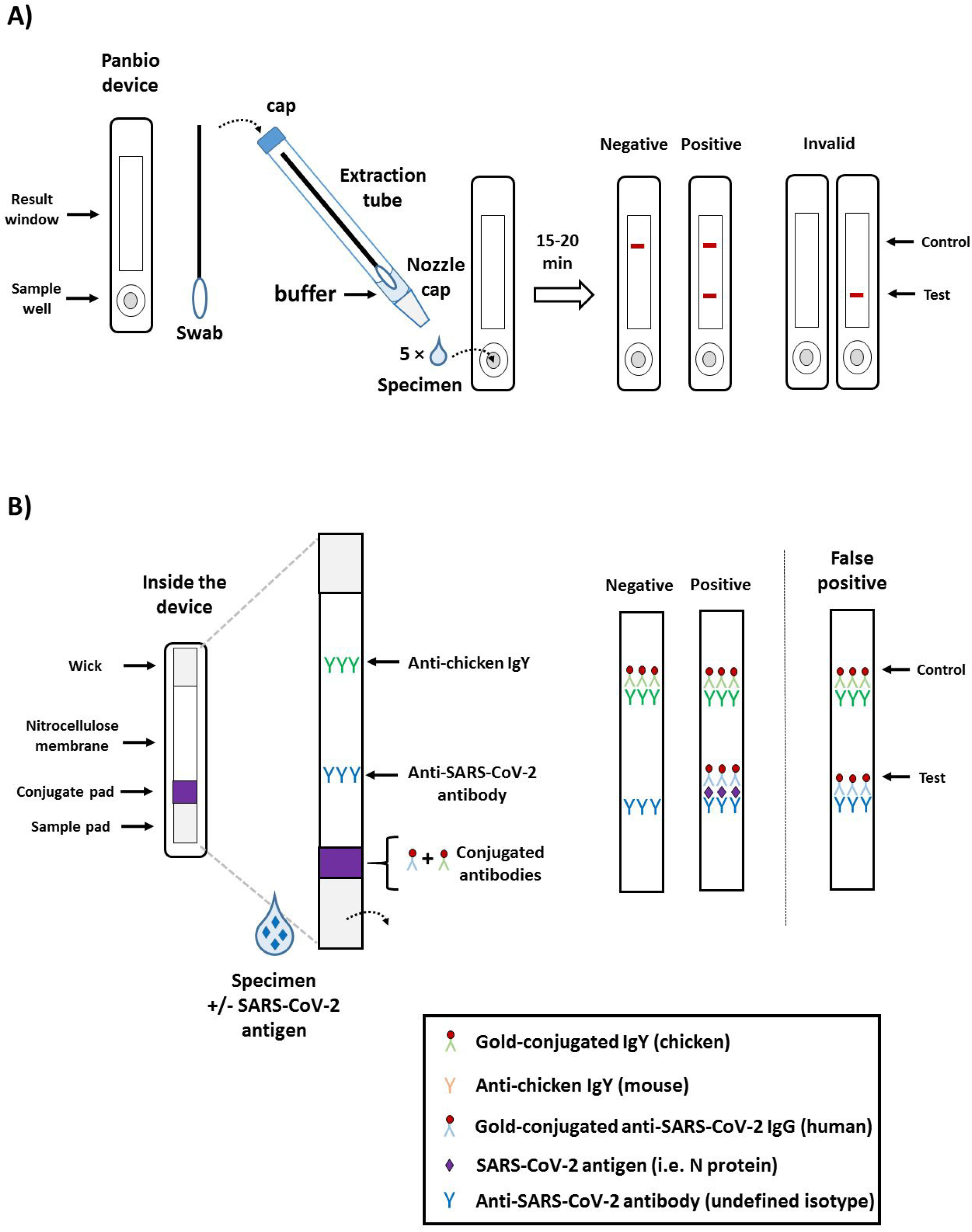
Summary and principle of the Panbio COVID-19 Ag Rapid Test Device. A) Panbio kit components and summary of the test procedure. The Panbio kit is designed for detection of SARS-CoV-2 antigen (i.e. nucleocapsid protein, or N protein). Following specimen collection, the swab is placed into an extraction tube pre-filled with 11-12 drops, or 300 µl) of buffer, and the tube cap is added. The tube is pinched to help extract the respiratory secretions from the swab, which in turn is rotated into the buffer. The nozzle cap is removed from the extraction tube, and 5 drops are placed into the sample well of the Panbio lateral flow device. After 15-20 minutes, the results are read and interpreted as depicted. B) The principle of the Panbio Ag-RDT relies on a nitrocellulose membrane pre-coated with anti-chicken IgY at the control line, and an anti-SARS-CoV-2 specific antibody at the test line. When the buffer/specimen solution is added to the sample well, the liquid flows progressively through the device using capillary action be sequentially flowing through the sample pad, the conjugate pad, the nitrocellulose membrane, and eventually into the wick. As the liquid comes into contact with the conjugate pad, both conjugated antibodies are resuspended (i.e. the gold-conjugated chicken IgY and the gold-conjugated human IgG specific to SARS-CoV-2). In absence of SARS-CoV-2 antigen (i.e. N protein), the conjugated anti-SARS-CoV-2 antibody will not interact with the anti-SARS-CoV-2 capture antibody at the test line; however, the conjugated chicken IgY will be captured by the anti-chicken IgY immobilized at the control line. This generates a red-colored band that can be visualized. In presence of SARS-CoV-2 antigen, a similar reaction occurs at the test line due to the interaction between the antigen and the conjugated and capture anti-SARS-CoV-2 antibodies. As seen in this study, false positives can occur with testing conditions falling outside of the manufacturer instructions, depicted here as non-specific interactions between the conjugated and capture anti-SARS-CoV-2 antibodies in absence of antigen.

### Assessing the role of the Panbio buffer and its components

PCR-grade water (Invitrogen) was chosen as a representative matrix to generate false positive Panbio results (Table 1). To assess the assay tolerability to buffer dilution, Panbio buffer was subjected to 2-fold serial dilutions in PCR-grade water, and testing was performed at 4°C, 20°C, and 37°C (Figure 2A). The exact composition of Panbio buffer is proprietary, yet according to the product insert, it consists of tricine, sodium chloride (NaCl), tween-20, proclin 300, and sodium azide (<0.1%). To assess the role of these components, the buffer was reverse engineered. Solutions of tricine (1 mM to 1M), from pH 3 to 12 were prepared (Figure 2B), with or without 1% tween-20. The contribution of ionic strength was assessed using NaCl (1 mM to 1M) in 100 mM tricine solutions pH 3 to 12 (Figure 2C).

**Figure 2.**
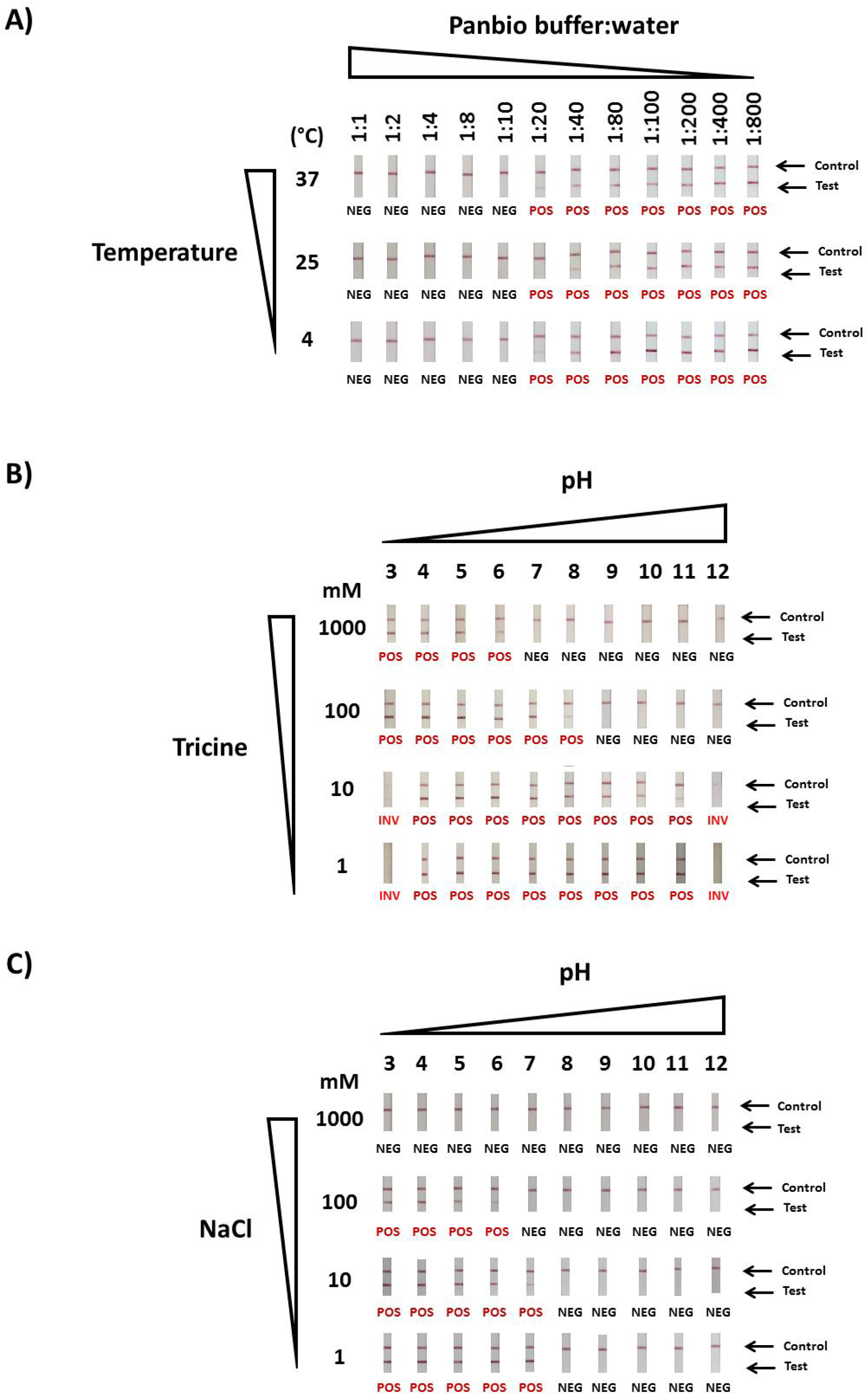
False positives SARS-CoV-2 Ag-RDT results can occur from Panbio buffer absence, dilution, or alterations. A) Artifact generation by Panbio buffer dilution in PCR-grade water at different temperatures (4°C, 25°C, and 37°C). B) False positives SARS-CoV-2 Ag-RDT results occurrence from uncontrolled pH and buffering conditions, or C) from changes in ionic strength from NaCl. All experiments were performed in absence of SARS-CoV-2 antigen. False positive reactions are indicated as POS (in red) under each Ag-RDT result.

### Effect of temperature and humidity on Panbio performance

According to manufacturer specifications, PanBio kits should be stored between 2 to 30 °C, and all kit components brought to room temperature (15 to 30 °C) for 30 minutes prior to use. To assess the impact of storage temperature, sealed PanBio test devices were incubated for one hour at 4°C, 20°C or 45°C (Table S1). The 45°C incubations were performed with 90% relative humidity (RH) using a Binder Constant Climate Chamber (model KBF 115) (Table S1). Test devices were removed from their packaging, and incubations were repeated under the same conditions. Testing was performed using 20 µL of gamma-irradiated SARS-CoV-2 into 280 µL of PanBio buffer. Viral stocks [at 1.2 ⨯ 10^6^ plaque forming units (PFU)/mL] were diluted in PBS (pH = 7.4) to concentrations spanning 1.2⨯10^5^ to 1.1 ⨯ 10^3^ PFU/mL (Table S1). PanBio buffer was used as negative controls. Freeze-thaw effects were investigated by incubation of test components at -20°C for 16 hours before thawing and testing.

### Investigations into possible causes of false positive results

Using “conjugate pad transplantation” (Figure 3A), the proprietary gold conjugated-antibodies of the Panbio device (i.e. the SARS-CoV-specific human IgG and the chicken IgY used for the control) were accessed from disassembled Panbio cassettes. Each conjugate pad was resuspended with 100 µl of Panbio buffer, PCR-grade water, or tricine solutions, and the suspensions were subjected to various treatments. Proteinase K (PK) (Qiagen GmbH., Hilden, Germany) was used at 100 µg/reaction for one hour at 56°C, followed by enzyme inactivation at 70°C for 10 minutes (Figure 3B). Untreated and heat treatment controls were included as controls (Figure 3B). The remaining conjugate-free pads are washed three times with 1 ml of water or buffer, dried using a Whatman #1 filter, and re-introduced into the Panbio cassettes. For testing, 25 µl of each water- or buffer-derived conjugated-antibody suspension was added onto the conjugate pads of reassembled cassettes, followed by addition of 5 drops into the sample well of either positive or negative controls processed in water or the kit buffer.

**Figure 3.**
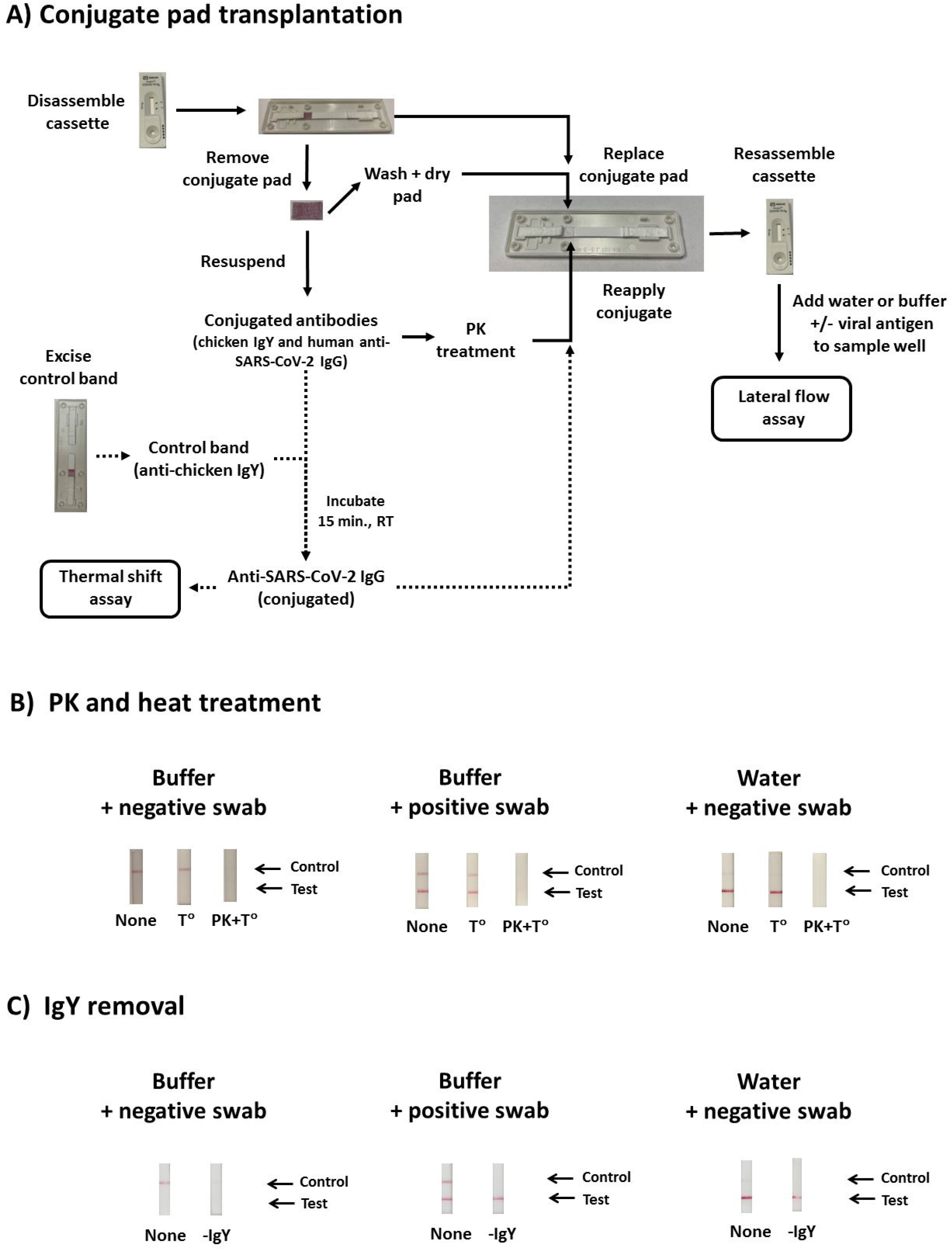
Impact of proteinase K (PK) and heat treatment on the conjugated SARS-CoV-2 antibody. A) Conjugate pad transplantation was used to access and investigate properties of the proprietary Panbio conjugated antibodies. Each step was followed to as depicted leading to treatment of the conjugated antibodies with proteinase K (PK) or heat (T°C), and comparisons were made with untreated controls (none). In some experiments (dashed arrows), the gold-conjugated antibody suspensions were pre-treated with mouse anti-chicken IgY (obtained from a fragment of the nitrocellulose membrane at the control line) to purify the gold-conjugated human IgG specific for SARS-CoV-2 conjugated antibody. This suspension was used for subsequent lateral flow and thermal shift assays. B) PK and heat treatments of the conjugated antibodies. Using conjugate pad transplantation, gold-conjugated antibody suspensions in Panbio buffer or water were treated for an hour with PK at 56°C, followed by heat inactivation of PK at 70°C for 10 min. Following re-introduction into Panbio cassettes of conjugated antibodies that were untreated (none), heat-treated (T°C), or PK-treated, and water was inoculated in the sample well. C) Removal of the conjugated chicken IgY from the conjugated antibody suspensions, to purify the conjugated SARS-CoV-2-specific antibody. Untreated (none) or pre-treatment (-IgY) are depicted for reassembled Panbio cassettes containing the purified conjugated SARS-CoV-2-specific antibody, which was then inoculated with PCR-grade water. For B) and C), similar reactions as performed for water were undertaken with positive or negative control swab, to demonstrate the method did not impact conjugate antibody function.

In a second set of experiments (Figure 3A, dashed lines), the control chicken IgY was removed from the SARS-CoV-2-specific IgG by pre-treatment of the conjugate suspensions with a fragment of the nitrocellulose membrane from the Panbio test device containing the immobilized mouse monoclonal anti-chicken IgY. Fragments were excised at approximately 3 mm on each side of the control line indicated on the Panbio cassette. For each 100 µl of conjugate suspension, one fragment was added, followed by a 15 min incubation at room temperature. Then, SARS-CoV-2-specific conjugated antibody were removed and subjected to lateral flow and thermal shift assays to explore possible pH-induced conformational changes (Figure 4).

**Figure 4.**
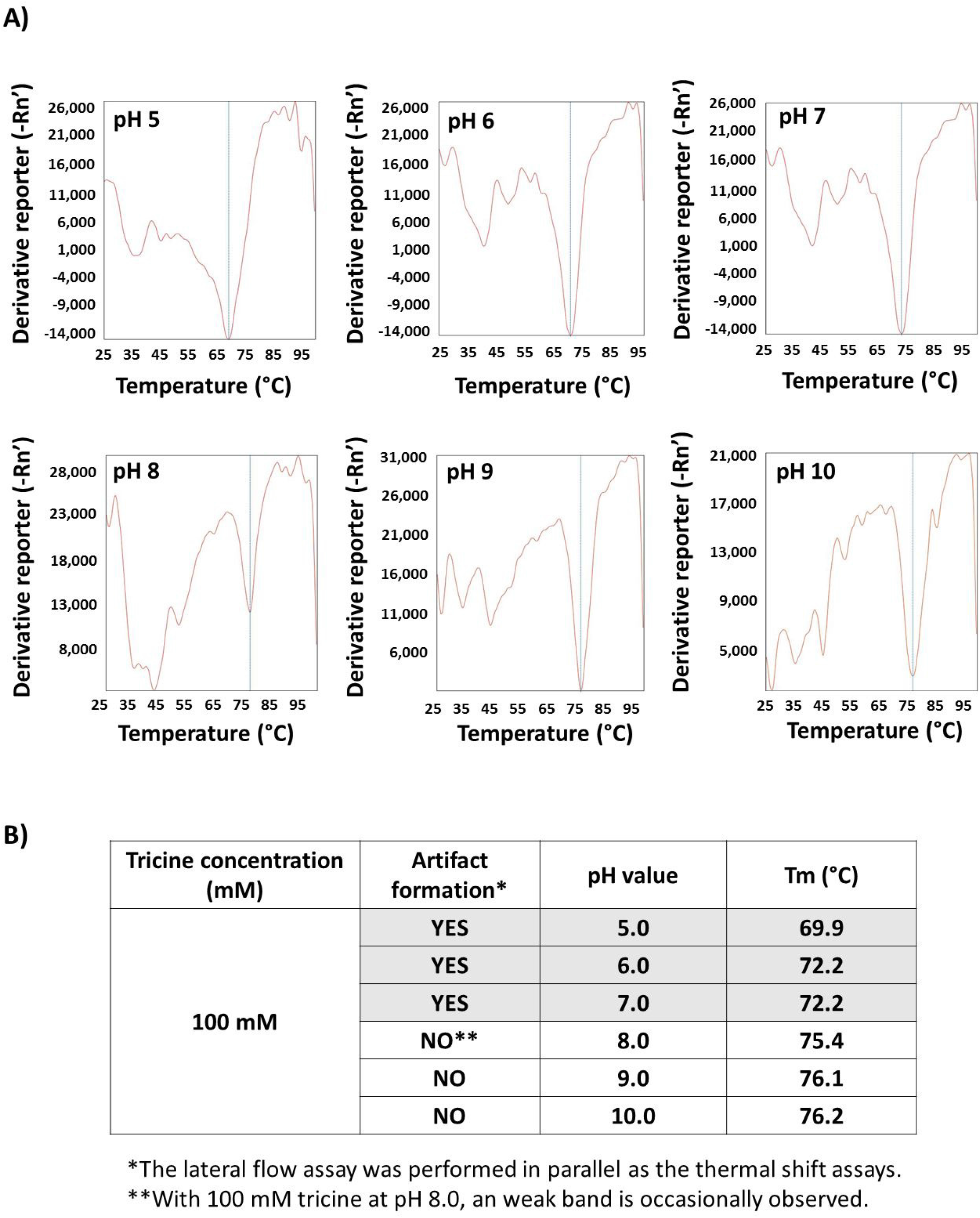
Thermal shift profiles for the Panbio gold-conjugated human IgG specific to SARS-CoV-2 at different pH values. All reactions were performed in 100 mM tricine, and representative thermal shift profiles are presented in A). Melting temperatures (Tm) changes based on pH are summarized in B), as well as a summary of which conditions were consistent (i.e. pH 5 to 7) or inconsistent (i.e. pH 8-10) with generation of false positive results when tested by Ag-RDT.

### Conjugated SARS-CoV-2 IgG thermal shift assays

Differential scanning fluorometry (DSF), also known as thermal shift assays, relies on monitoring temperature-dependent unfolding of a protein, in presence of a fluorescent dye that is quenched in water but fluoresces when bound to hydrophobic residues.[22-24] As a native protein is unfolded with heat, different hydrophobic residues are exposed, and the melting temperature (Tm) can be calculated for various test conditions. In this study, 25 μl reaction containing 10× SYPRO Orange (Invitrogen, Eugene, Oregon, USA) was added to the gold-conjugated human IgG specific to SARS-CoV-2 resuspended in Panbio buffer or 100 mM of tricine at pH values consistent (i.e. pH 5-7) and inconsistent (i.e. pH 8-10) with artifact formation (Figure 4A and B). A similar set of experiments with performed with addition of 1% tween-20. Melting curve analysis was performed by increasing the temperature from 25°C to 99.9°C at a ramp rate of 1% with continuous fluorescence at 610 nm using an Applied Biosystems 7500 Fast instrument. Tm values were calculated by manufacturer software (Figure 4B).

## Results

### False positives in food, water, buffers, media, and clinical specimens

With the exception of soft drinks and some milk products with high fat content which produced negative or weak false positive reactions, most of the food products that were tested directly onto the Panbio cassette produced a strong positive SARS-CoV-2 signals that resembled those obtained with the kit positive control (Table 1). It is interesting that, milk products are often used as blocking agents in immunoassays to prevent non-specific binding of antibodies.[1,27] Direct testing of known highly acidic samples caused invalid results for both Panbio and Veritor. All other products were Veritor-negative. When nasal swabs were used to sample the various products and processing occurred with manufacturer buffer, no false positives or invalid results were observed.

Multiple water samples were evaluated with tested pH values between 4.00 and 9.33, and differences in supplier-described purification methods, and mineral and electrolyte composition (Table 1). Direct testing onto Panbio test devices showed strong false positive SARS-CoV-2 signals, while samples diluted in Panbio buffer did not produce any artifacts. Notably, water samples near the pH of the Panbio buffer (pH 8.78) also displayed strong false positive signals, suggesting the mechanism behind artifact formation is not, or not solely, pH-dependent. To investigate the possible roles of buffering capacity and ionic strength, commonly used laboratory buffers and buffer-containing viral transport media spanning various pH values (5.62 to 8.78) were tested (Table 1). With the exception of Tris-EDTA (TE), all other buffers and media generated weakly positive or negative results (Table 1). All water samples, buffers, and media were RT-PCR and Veritor-negative, suggesting absence of viral RNA and nucleocapsid antigen, respectively (Table 1).

Given that weak false positive were results observed with UTM, PBS, and saline, direct testing was performed on clinical specimens containing these media and buffers. With direct testing onto Panbio cassettes, false positive results were seen in 93.3% of NP swabs in UTM, 86.7% of OP/N swabs in PBS, 90.0% of BALs, and 90.0% of the saline gargles (Table 1). All specimens were negative when Panbio buffer was used, which was consistent with the Veritor and RT-PCR results.

### Role of the Panbio buffer and its components

Panbio buffer diluted in water at ratios greater than 1:8, and occasionally at 1:10, resulted in artifact formation (Figure 2A). Similarly, when buffering capacity was poor or lost when using low tricine concentrations (1 or 10 mM), strong false positive signals were seen across a broad range of pH values (Figure 2B). In contrast, high tricine concentrations (100 mM or 1M) prevented artifact formation at pH 9 and above, which is consistent with the measured pH of Panbio buffer at 8.78 (Figure 2B and Table 1). Similar to the buffering capacity, regulated ionic strength also played an important role, as high NaCl concentrations (100 mM or 1M) reduced or prevented false positive results, whereas lower concentrations (1 and 10 mM) mirrored NaCl-free conditions (Figures 2B and 2C). Invalid results sometimes obtained at pH 3 and 12 (Figure 2B and 2C). Tween-20 (1%) was added to all tricine solutions, but had no impact on results (data not shown). Antimicrobial agents in the Panbio buffer (i.e. proclin 300 and sodium azide) were not investigated due to their unlikely contribution to artifact generation.

### Investigations into the mechanism of artifact generation

Following conjugate pad transplantation (Figure 3A), positive and negative control swabs displayed expected results after inoculation onto re-assembled Panbio cassettes in which resuspended conjugated antibodies were re-introduced. Water-resuspended conjugated antibody generated a strong false positive target signal, which was eliminated following PK treatment (Figure 3B). Removal of the gold-conjugated IgY antibody from the conjugate suspensions did not impair Panbio test performance, and the strong false positive SARS-CoV-2 signal from water remained (Figure 3C). These findings suggest that the gold-conjugated human anti-SARS-CoV-2 IgG is responsible for the non-specific interactions with the immobilized anti-SARS-CoV-2 capture antibody on the test device nitrocellulose membrane. Thermal shift assays in 100 mM tricine solutions were used to compare structural differences of the anti-SARS-CoV-2 IgG at pH values consistent (i.e. pH 5-7) or inconsistent (i.e. pH 8-10) with artifact formation (Figure 4A and B). Tm values were significantly different in tricine solutions between pH 5 to 7 (at 68.4 ± 2.6, 71.4 ± 1.2, 72.5 ± 1.0, respectively) compared to pH 8 to 10 (at 75.6 ± 1.0, 76.8 ± 1.1 and 77.6 ± 1.4, respectively) (Figure 4A and B). Tm values at pH 4 and 11 were inconsistent, while no Tm values could be established at pH 3 and 12. Of note, the Panbio buffer could not be used directly for thermal shift experiments, due to high background fluorescence with SYPRO orange. The cause of this background fluorescence was revealed in tricine solutions containing 1% Tween-20, which demonstrated similar interference.

### Impact of heat and humidity on Panbio kit function

In all test conditions evaluated (Table S1), no deleterious effects on test sensitivity or specificity were observed concerning temperature or humidity. In a complimentary series of experiments, Panbio buffer dilutions showed similar findings, regardless of operating temperature (Figure 2A).

## Discussion

False positive and negative results occur with any diagnostic test, but are increasingly likely when manufacturer recommendations are not followed.[1,13,14] This study demonstrated in absence of manufacturer buffer, a variety of food, water, laboratory buffer, specimen transport media, and clinical specimens resulted in false positive reactions with the Panbio Ag-RDT. These data are consistent with others [25] who recognized the importance of Ag-RDT kit buffers. On the other hand, false negative results would typically be expected with aberrant test conditions. The generation of false positive signals demonstrated in this study prompted an investigation into the underlying causes of this phenomenon. Uncontrolled conditions of pH, buffering capacity, and ionic strength, all favored artifact generation, whereas temperature and humidity were not contributory under the tested parameters. In review of the literature, possible causes of false positive Ag-RDT results include cross-reactions [26], interfering substances [27], and improper operating or storage conditions for temperature or humidity.[28]. Cross-reacting or interfering substances common to all samples tested in the study is unlikely. Temperature extremes have been shown to induce conformational changes in SARS-CoV-2 antibodies, leading to non-specific binding [29]; however, in this study, Panbio was unaffected by temperature and humidity conditions evaluated. As described below, the most likely cause of Panbio false positive results was aberrant protein-protein interactions faced with improper buffer conditions, ionic strength, or pH.

In a previous study [25], 20 of 27 of the malaria Ag-RDTs brands evaluated showed false positive reactions when the manufacturer buffer was replaced with saline, tap water, or distilled water. Distilled water alone generated false positive reactions [25], similar to what was observed in this study with Panbio (Table 1). Possible explanations for their findings included inefficient resuspension of blocking agents, altered capillary flow rates and decreased flushing of contaminating substances, and finally, non-specific interactions between the conjugated and capture antibodies faced with uncontrolled buffering and ionic strength conditions.[25] Tricine is a zwitterionic amino acid with a pKa of 8.26, and would be negatively charged at the measured pH of 8.78 in the Panbio buffer. Therefore, under recommended testing conditions, tricine may mask positively charged residues on the SARS-CoV-2-specific conjugated and capture antibodies, while uncontrolled buffer conditions would favor aberrant electrostatic or hydrophobic interactions between the two antibodies, resulting in false positive results. Supporting this theory, PK treatment eliminated the false positive Panbio results generated by water, and the propensity to generate this artifact varied with buffering capacity, pH, and ionic strength. Removal of the gold-conjugated chicken IgY (used for control band detection) did not alter formation of the SARS-CoV-2 target artifact formation, suggesting the conjugated SARS-CoV-2-specific IgG alone is responsible for artifact formation through non-specific binding to the SARS-CoV-2-specific capture antibody immobilized on the nitrocellulose membrane (Figure 1B). Finally, thermal shift assays were performed on the conjugated anti-SARS-CoV-2 IgG, and pH-dependent conformation changes were observed under conditions causing false positive results or not (Figure 4). This study was not able to further investigate pH-dependent binding interactions between the conjugated and capture anti-SARS-CoV-2 antibodies, as the latter is immobilised on the nitrocellulose membrane, and not available in an unfixed formulation due to the proprietary nature of the Panbio assay.

False positive reactions with the Panbio Ag-RDT have the potential to cause a significant impact to Public Health. To date, over 200 million Panbio Ag-RDT tests have been distributed to over 120 countries worldwide, for use in healthcare settings, businesses, or home self-testing. In low prevalence populations, positive Ag-RDTs are typically confirmed by clinical laboratories with NAATs, thereby limiting the overall public health impact of the possible artifacts described in this study.[1-3] However, in programs where home self-testing kits are deployed [20,21,30], it is important to educate users on the importance of strict adherence to manufacturer instructions. Another area for consideration is outdoor testing strategies (e.g. drive-thru testing), where the Panbio kit supplies may be exposed to precipitation and fluctuations in temperature and humidity.[30] While temperature and humidity did not alter the Panbio performance in this study, rain water was shown to cause false positive reactions if processed without buffer. Haage et al. [28] demonstrated that prolonged exposure to elevated temperatures affected the sensitivity of SARS-CoV-2 detection by some Ag-RDTs, whereas low temperatures impaired specificity of assays including Panbio.[25] However, an alternative explanation for the false positives observed at low temperatures by these investigators could be the use non-validated specimen types (i.e. NP swabs in PBS).[25] In this study, PBS alone caused false positive results in absence of buffer. The quantity of PBS material (i.e. 20 µl in approximately 300 µl of buffer) used by Haage et al. [25] was similar to the limit of tolerability of Panbio to dilution in water observed in this study of between 1:8 to 1:10.

It should be noted that the findings of this study with false positives results observed with Panbio when tested outside manufacturer claims should not be extrapolated to other Ag-RDTs without supporting evidence, as a second SARS-CoV-2 antigen test (i.e. BD Veritor) did not show similar findings. Other Ag-RDTs could rely on different assay principles, and would need to be investigated independently.

Overall, this manuscript provides rigorous scientific evidence that erroneous false positive SARS-CoV-2 results can occur with improper test conditions with the Panbio Ag-RDT, resulting in non-specific interaction between the SARS-CoV-2-specific conjugated and capture antibodies. While generation of false positive results from direct testing of products onto Panbio Ag-RDT devices may not surprise some healthcare professionals, having a better understanding of the importance of the buffer and its components, as well as knowing the mechanism of false positive generation, can help dispute unsound demonstrations on social media, and help inform users on the value of following the manufacturer instructions.

## Data Availability

All data has been included in the manuscript or supplemental materials.

## Acknowledgement

The authors would like to thank the Special Pathogens Program of the National Microbiology Laboratory (NML) (Winnipeg, MB) for the gamma-irradiated SARS-CoV-2 virus used in this study. We would also like to recognize the ongoing efforts of the all the NML and NSH staff for their dedication and exceptional services throughout the pandemic, including the help with RT-PCR testing during this evaluation. The authors would also like to thank all the volunteers and healthcare professionals who dedicate their time at popup clinics and other settings that use Ag-RDTs or NAATs. You are all instrumental for our pandemic responses, and safety in our communities.

## Author contributions

JH, AB, NS, and JR identified this phenomenon, and were involved in food sample testing. JL, RD, GP, and TH designed and undertook testing of water, buffer, media, and gargles. JL designed and performed the buffer reverse engineering, conjugate pad transplantation, PK experiments and DSF experiments. MB, AM, EM, BH, and PS designed and performed the temperature and humidity experiments. GP, JL, and TH wrote the initial draft of the manuscript, with all authors contributing to the final version.

## Transparency declaration

The authors declare that they have no conflicts of interest. This work received no private or public funding, with the exception of the Panbio kits that were provided in-kind from the Public Health Agency of Canada (PHAC).

## Ethics

This evaluation was deemed exempt from Nova Scotia Health Research Ethics Board approval, as the activities described in this manuscript were conducted in fulfillment of ongoing verification of SARS-CoV-2 diagnostic assays used in Nova Scotia, and are therefore considered a quality assurance initiative. Clinical specimens tested were obtained from anonymized residual samples collected for routine diagnostic testing for SARS-CoV-2 from consenting participants, and all data related to clinical specimens were provided anonymized, de-identified, and were used solely with the intent to evaluate the potential for false positives in these clinical specimen types for rapid antigen testing programs used in Nova Scotia.

