## Supplemental Table S1 for "Generation of false positive SARS-CoV-2 antigen results with testing conditions outside manufacturer recommendations: A scientific approach to pandemic misinformation"

**Table S1.** Impact of temperature and humidity on Panbio test performance.

| Temperature (°C) | | Results*** | | | | | | |
|---|---|---|---|---|---|---|---|---|
| | | | SARS-CoV-2 concentration (PFU/ml) | | | | | |
| **Pre-treatment*** | **Test Conditions**** | **PanBio Buffer** | **$1.2x10^5$** | **$1.2x10^4$** | **$5.0x10^3$** | **$2.5x10^3$** | **$1.25x10^3$** | **$1.1x10^3$** |
| **20** | **20** | 0/3 | 3/3 | 3/3 | 3/3 | 3/3 | 3/3 | 0/3 |
| | **4** | 0/3 | 3/3 | 3/3 | 3/3 | 3/3 | 3/3 | 0/3 |
| | **45** | 0/3 | 3/3 | 3/3 | 3/3 | 3/3 | 3/3 | 0/3 |
| **45** | **20** | 0/3 | 3/3 | 3/3 | 3/3 | 3/3 | 3/3 | 2/3 |
| | **4** | 0/3 | 3/3 | 3/3 | 3/3 | 3/3 | 3/3 | 1/3 |
| | **45** | 0/3 | 3/3 | 3/3 | 3/3 | 3/3 | 3/3 | 0/3 |
| **4** | **20** | 0/3 | 3/3 | 3/3 | 3/3 | 3/3 | 3/3 | 1/3 |
| | **4** | 0/3 | 3/3 | 3/3 | 3/3 | 3/3 | 3/3 | 1/3 |
| | **45** | 0/3 | 3/3 | 3/3 | 3/3 | 3/3 | 3/3 | 0/3 |
| **-20** | **20** | 0/3 | N/A | N/A | 3/3 | 3/3 | 3/3 | 0/3 |

*Pre-treatment was performed with test components in packaging.

**Samples stored at 45°C were incubated at a relative humidity of 90-100%

*** Results are the summary obtained from triplicate testing.
